# Diagnostic performance of LumiraDx SARS-CoV-2 rapid antigen test compared to PCR for diagnosis of COVID-19 in Liberia

**DOI:** 10.1101/2023.08.01.23293521

**Authors:** Moses Badio, Christina Lindan, Kumblytee Johnson, Cavan Reilly, Julie Blie, Katy Shaw-Saliba, Tamba Fayiah, John McCullough, J. Tanu Duworko, Elizabeth Higgs, Jeffrey Martin, Lisa Hensley

## Abstract

**Background:** Diagnosis of SARS-CoV-2 infection in Liberia has primarily relied on polymerase chain reaction (PCR)-based testing at the country’s National Reference Laboratory. This centralized approach caused reporting delays, prompting evaluation of point-of-care antigen-based tests. We assessed the test performance of the LumiraDx™ SARS-CoV-2 Ag test (LumiraDx™, London, UK) in this setting.

**Methods:** We tested ambulatory individuals screened for enrollment into an observational cohort study of COVID-19 sequelae in Monrovia in 2021. We compared the results of LumiraDx testing of anterior nasal swab specimens to the results of PCR BioFire® R2.1P (bioMérieux, Salt Lake City, Utah) on eluent from nasopharyngeal swabs.

**Results:** We evaluated 348 individuals. Among the 274 persons with symptoms, 49.3% were PCR-positive and 36.5% were antigen-positive. The sensitivity, specificity, negative predictive value (NPV), and positive predictive value (PPV) of LumiraDx in this group were 72.6% (95% CI: 64.3%-79.9%), 98.6% (95% CI: 94.9%-99.8%), 78.7% (CI: 71.9%-84.6%), and 98.0% (CI: 93.0%-99.8%), respectively. Among the 74 asymptomatic individuals, 12.2% were positive by PCR, and 5.5% by antigen testing, resulting in a sensitivity, specificity, NPV, and PPV of LumiraDx of 44.4% (95% CI: 13.7%-78.8%), 100% (95% CI: 94.5%-100%), 92.9% (CI: 84.1%-97.6%), and 100% (CI: 39.8%-100%), respectively.

**Conclusion:** Although the specificity and PPV of LumiraDx for diagnosing SARS-CoV-2 were high among persons with and without symptoms, the sensitivity was unacceptably low in both groups, and much less than that reported by the manufacturer. Before new point-of-care diagnostics are adopted, test performance needs to be assessed in the local setting.

## Introduction

A global shortage of laboratory tests to detect SARS-CoV-2 infection occurred during the beginning of the COVID-19 pandemic due to high demand and disruptions in supply chains; this was particularly true in many parts of sub-Saharan Africa (SSA)[1-4]. In resource-limited settings, identifying persons with COVID-19 was further complicated by the lack of alternative and accurate rapid tests, causing reliance on reverse-transcriptate polymerase chain reaction (PCR) to detect the presence of SARS-CoV-2 RNA. PCR technology has been the gold standard for diagnosis, but requires specialized and costly equipment and reagents; processing can result in lengthy delays in obtaining results[2], hindering recommendations for people to quarantine or isolate, and preventing timely provision of available treatment, in turn impairing the ability to reduce ongoing transmission[3]. The limitations of laboratory-based PCR testing spurred the development of point-of-care (POC) rapid antigen-based diagnostics, well as self-administered home-based testing[4].

In Liberia, laboratory-confirmed diagnosis of SARS-CoV-2 infection relied exclusively on PCR testing at the National Reference Laboratory (NRL). Specimens from all 15 counties in the country had to be transported to the NRL; the laboratory was difficult to access and could only be reached using unpaved roads. This centralized approach caused reporting delays of at least two weeks due to low throughput of the PCR platform and the logistical difficulty of transporting specimens. These constraints were acutely experienced during a surge of SARS-CoV-2 transmission from June to August 2021, primarily attributed to the delta-variant[5,6]. During this period, a large number of people with respiratory symptoms presented to health facilities, but diagnosis and management of COVID-19 were impaired due to the inability to obtain test results quickly. Neither POC nor self-administered home-based rapid antigen tests were available in Liberia at the time.

The PREVAIL COVID-19 Observational Study (PCOS), a research collaboration between the U.S. National Institute of Allergy and Infectious Diseases/National Institute of Health (NIAID/NIH) and the Liberian Ministry of Health (MOH), was set up in August 2020 at the JFK Hospital, the main referral hospital in the country. The goals of PCOS were to evaluate the acute and long-term sequelae of SARS-CoV-2 infection by evaluating symptomatic and asymptomatic infected individuals over time, as well as a comparator group of uninfected persons. Potential participants were screened for infection using the BioFire® R2.1P (bioMérieux, Salt Lake City, Utah) at the PCOS research laboratory at the JFK hospital. The Liberian MOH requested assistance from PCOS to evaluate the utility and accuracy of the LumiraDx™ SARS-CoV-2 antigen test (LumiraDx, London, UK). This platform was being considered by the MOH for use at clinical sites around the country to hasten diagnoses and reduce the burden on NRL, if it performed well in comparison to PCR. Assessment of the local test performance of LumiraDx was considered important, in part because it was not known how POC diagnostic tests would perform in the Liberian population, especially given early indications of less severe clinical disease in many parts of SSA compared to populations in Europe, South America, and the U.S[7-9].

## Methods

### Study design and summary

We evaluated the test performance of the LumiraDx antigen test compared to the results of the BioFire PCR on eluent from nasal swabs obtained from symptomatic and asymptomatic individuals being screened for enrollment into PCOS. Both tests had received U.S. FDA Emergency Use Authorization at the time of the study [8,9].

### Study population

Persons of any age who were ambulatory and seeking outpatient care at the JFK Hospital for any reason and who had symptoms consistent with COVID-19 were referred by clinicians to the PCOS testing site for study information, testing for SARS-CoV-2, and recruitment. Individuals from the community who voluntarily sought testing, regardless of symptoms, could also be tested, and if interested, evaluated for enrollment. In this analysis, we present data from a consecutive sample of persons who were screened from June to August 2021, regardless of whether they subsequently enrolled in PCOS. All persons provided written informed consent for screening procedures, and a completed a separate signed consent if they decided to enroll in PCOS. The study was reviewed and approved by the Liberian National Research Ethics Board.

### Measurements and laboratory testing

Participants completed a short interviewer-administered survey about socio-demographics and whether they had experienced any of 10 COVID-19-related symptoms during the last two weeks; at the time, these symptoms were included on the MOH screening algorithem for COVID-19.

Nasopharyngeal (NP) HydraFlock swabs for PCR testing and anterior nasal HydraFlock swabs for antigen testing were collected by trained laboratory technicians. Each swab was rotated for 10-15 seconds in both nares. NP swabs were immediately placed in 3 mL of viral transport media, and eluent tested within 45 minutes using BioFire at the PCOS research laboratory co-located at JFK Hospital, following manufacturer guidelines[11]. The BioFire targets the viral spike (S) and membrane (M) proteins.Anterior nasal swabs were immediately placed in pre-packaged extraction buffer and tested within 30 minutes using the LumiraDx instrument according to the manufacturer’s instructions[12]. This test uses a microfluidic immunofluorescence-based assay for the qualitative detection of the SARS-CoV-2 nucleocapsid protein. Laboratory technicians wore personal protective equipment (PPE) including gowns, surgical caps, masks, and gloves during specimen collection, handling, and testing; gloves were changed between the processing of each individual specimen.

### Statistical analysis

The distribution of age, gender, and symptoms were computed using proportions. We determined the ‘prevalence’ or pre-test probabitiy of infection in the whole sample, and among persons with and withought symptoms, based on the proportion with a positive BioFire PCR test. We calculated the sensitivity, specificity, positive predictive value (PPV), and negative predictive value (NPV) of LumiraDx with 95% confidence intervals (CI) among the entire sample, as well as stratified by whether persons had symptoms or not. Based on the sensitivity and specificity of the antigen test in our sample, we estimated the PPV and NPV among persons with and without symptoms at varying levels of pre-test probability of SARS-CoV-2 infection. Analyses were conducted using Stata version 17.0 (Stata Corporation, College Station, Texas, United States).

## Results

Among the 348 individuals evaluated, 44.3% were female; the age range was 8 to 86 years, and 44.3% were 30 years of age or less (Table 1). More than three-fourths (78.7%) reported at least one COVID-19-related symptom in the last two weeks. The most common symptoms were weakness (50.9%), cough (42.2%), body pain (37. 9%), and fever (37.1%).

**Table 1.** Demographic characteristics and symptoms of participants tested for SARS-CoV-2 infection with LumiraDx™ antigen test and BioFire® PCR, Monrovia, Liberia, June – August 2021.

|  | **N** | **%** |
| --- | --- | --- |
| **All** | **348** | **100** |
| **Age<sup>1</sup>, years** |  |  |
| ≤30 | 152 | 44.3% |
| 31-50 | 63 | 18.4% |
| >50 | 128 | 37.3% |
| **Female** | 154 | 44.3% |
| **Symptomatic** |  |  |
| Any symptom | 274 | 78.7% |
| **Symptoms<sup>2</sup>** |  |  |
| Fever | 129 | 37.1% |
| Cough | 147 | 42.2% |
| Sneezing | 56 | 16.1% |
| Sore throat | 40 | 11.5% |
| Shortness of breath | 121 | 34.8% |
| Absence of smell | 66 | 19.0% |
| Weakness/fatigue | 177 | 50.9% |
| Body pain | 132 | 37.9% |
| Headache | 123 | 35.3% |
| Diarrhea | 33 | 9.5% |

<sup>1</sup>Small differences in N are due to missing data
<sup>2</sup>Only among the 274 who had at least one symptom

Among all NP specimens tested by BioFire, 41.4% were positive for detection of SARS-CoV-2 RNA; among all anterior nasal specimens tested using LumiraDX, 29.9% were positive (Table 2). Among samples from the 274 persons with symptoms, 49.3% were PCR-positive, and 36.5% were antigen-positive. The sensitivity, specificity, NPV and PPV of LumiraDx in this group were 72.6% (95% CI: 64.3% to 79.9%), 98.6% (95% CI: 94.9% to 99.8%), 78.7% (71.9%, 84.6%), and 98.0% (93.0%, 99.8%) respectively. Among samples from the 74 asymptomatic individuals, 12.2% were positive by PCR and 5.4% were positive by antigen-testing. The sensitivity, specificity, NPV, and PPV of LumiraDx in persons without symptoms were 44.4% (95% CI: 13.7% to 78.8%), 100% (95% CI: 94.5% to 100%), 92.9% (84.1%, 97.6%), and 100.0% (39.8%, 100%) respectively.

**Table 2.** LumiraDx™ SARS-CoV-2 antigen test performance compared to BioFire® R2.1P PCR among persons with and without COVID-19 symptoms, Monrovia, Liberia, June – August 2021.

| LumiraDx | BioFire |  |  | LumiraDx test performance | % | (95% CI) |
| --- | --- | --- | --- | --- | --- | --- |
|  | Positive | Negative | All |  |  |  |
| **All** |  |  |  | Sensitivity | 70.8% | [62.7%, 78.1%] |
| Positive | 102 | 2 | 104 | Specificity | 99.0% | [96.5%, 99.9%] |
| Negative | 42 | 202 | 244 | Positive predictive value | 98.1% | [93.2%, 99.8%] |
| Total | **144** | **204** | **348** | Negative predictive value | 82.8% | [77.5%, 87.3%] |
| **Symptomatic** |  |  |  | Sensitivity | 72.6% | [64.3%, 79.9%] |
| Positive | 98 | 2 | 100 | Specificity | 98.6% | [94.9%, 99.8%] |
| Negative | 37 | 137 | 174 | Positive predictive value | 98.0% | [93.0%, 99.8%] |
| Total | **135** | **139** | **274** | Negative predictive value | 78.7% | [71.9%, 84.6%] |
| **Asymptomatic** |  |  |  | Sensitivity | 44.4% | [13.7%, 78.8%] |
| Positive | 4 | 0 | 4 | Specificity | 100.0% | [94.5%, 100%] |
| Negative | 5 | 65 | 70 | Positive predictive value | 100.0% | [39.8%, 100%] |
| Total | **9** | **65** | **74** | Negative predictive value | 92.9% | [84.1%, 97.6%] |

Based on our calculated sensitivity and specificity, we estimated that among persons with symptoms, the PPV of LumiraDx would be >=90% when the true prevalence of infection is 15% or higher (Figure). In symptomatic persons, the NPV increases as the prior-probability of infection,decreases, reaching >= 90% when the true prevalence is 30% or less. Among asymptomatic persons, the NPV would be high, or >=90%, when the true prevalence of infection is 16% or less. The PPV among asymptomatic individuals at varying levels of prevalence could not be calculated, however, because the specificity was 100%.

**Figure 1.**
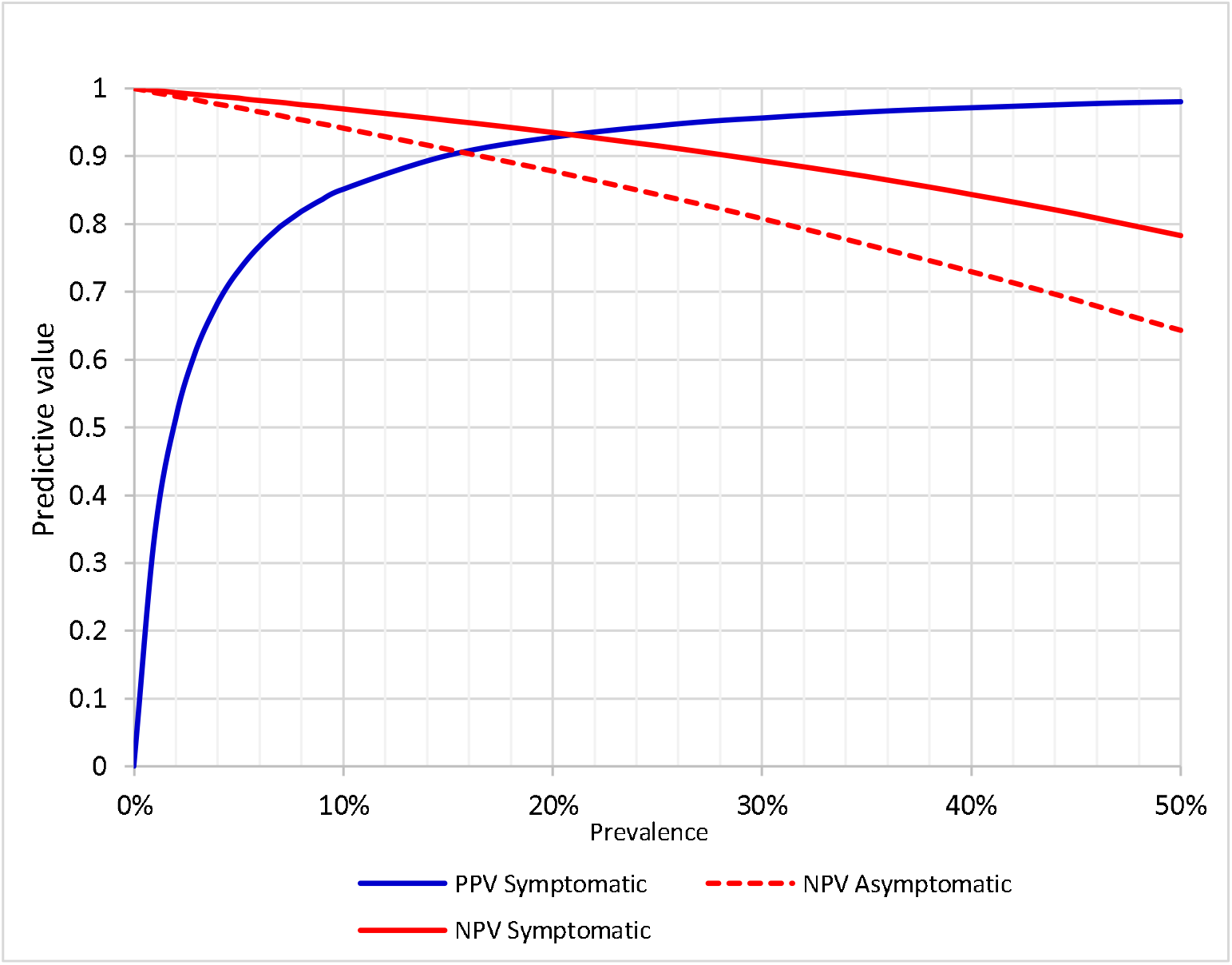
Predictive value of the LumiraDx™antigen test in populations with varying prevalence of acute SARS-CoV-2 infection, among symptomatic and asymptomatic individuals. Note: The specificity among asymptomatic individuals is 100%. Hence, the PPV cannot be calculated.

## Discussion

We evaluated the test performance of the LumiraDx SARS-CoV-2 Ag test compared to the BioFire PCR in Liberia and found that among persons both with and without symptoms, the antigen test had a high specificity and PPV of 98% -100%. In contrast, the sensitivity was unacceptably low: 73% in persons with symptoms, and 44% among asymptomatic people. This means that in someone presenting with symptoms suggestive of COVID-19, a negative test would need to be corroborated by a PCR or other nucleic acid amplification test (NAAT), particularly at higher levels of community prevalence. Because of these findings, the Liberian MOH decided not provide LumiraDx to local hospitals and clinics as an alternative decentralized method of diagnosing infections.

The sensitivity of LumiraDx that we found in our Liberian sample was substantially different from the 97.6% presented in the manufacturer’s insert [13] and a publication based on the same data [14]. It should be noted that the the product insert and the paper by Drain et al, do not specify the symptom status of those tested using anterior nasal swabs, or how participants were selected. Even if we assume all persons in those studies were symptomatic, the 73% sensitivity we found among persons with symptoms was still much lower in the product insert. It is possible that individuals in the manufacturer’s study had more severe COVID-19 disease compared to our participants, with a higher viral load and thereby more likely to be detected by an antigen test [13,14].

Several published studies have also evaluated the performance of LumiraDx compared to PCR as the gold standard. Similar to our findings, most found a high specficity among both symptomatic and asymptomac individuals [15-18]. Reported sensitivity varied widely, however. Some studies found sensitivities of 90% or greater [15,17,18], while others reported values closer to the 73% we observed [19,16,20]. It is worth noting that studies reporting a high sensitivity primarily tested patients in emergency departments or residents of nursing homes, who might have been sicker than our participants [15,18,17]. Studies that found a lower sensitivity generally evaluated individuals seeking care at outpatient departments, who may have been more similar our ambulatory sample [19,21].

The variability in test performance among studies and by patient population, highlights the importance of evaluating test diagnostics in the population and setting in which they are to be used. In addition, the probability that a negative or a positive test finding is accurate depends on the probability of true infection among persons being tested, which varies across geographic regions, and is another reason to determine the accuracy of a test where it will be used. It should be noted that the only other study from SSA that evaluated LumiraDx in addition to ours was performed in Mozambique, and found similarly low sensitivity as in our study, or 80% among those with symptoms, and 48% among those who were asymptomatic[21].

We do not believe that the low sensitivity we found was due to laboratory or other procedural errors. The on-site research laboratory used for many other PREVAIL studies in addition to PCOS, conducts verification and validation testing to ensure the accuracy and reliability of findings. However, our study did have several limitations. First, the sample size for asymptomatic individuals was small, resulting in wide confidence intervals around some estimates. Secondly, the manufacturer recommends that LumiraDx not be used more than 12 days after symptom onset. We did not collect information about when symptoms began; therefore, it is possible that the low sensitivity and NPV among symptomatic participants could partially be a results of including persons beyond the window of detection for the LumiraDx. Finally, we only asked participants about 10 possible COVID-19 related symptoms. Since then, many additional symptoms have been associated with acute SARS-CoV-2 infection [24]. Misclassifying some persons with COVID-19 symptoms that we did not recognize, could have resulted in us calculating a somewhat falsely low NPV.

The LumiraDx platform could be a useful test to quickly triage persons with a positive antigen test in clinical and resource limited settings when confirmatory NAAT testing is not rapidly available or affordable [22-24]. The turnaround time to obtain a test result once the sample has been inserted into the machine, is only 12 minutes, and use of the platform does not required skilled technicians. The main logistical drawbacks are that the platorm needs to be used in a laboratory setting, and only one sample can be evaluated at a time. Despite the several advantages of the LumiraDx diagnostic test, however, the low NPV means that a negative result in someome with symptoms can not be used to rule-out an infection, requiring a confirmatory test.

Our study demonstrated both the potential usefulness and limitations of using this antigen test in a country in west Africa, and highlighted the importance of not assuming pubished test performance is applicable across regions and different populations. The COVID-19 pandemic has waned, and the urgency of alternative screening and testing methods for SARS-CoV-2 infection is no longer acute. During the last several years, home-based and self-administered testing using convenient and rapid lateral flow tests have been widely used in many parts of the world. Unfortunately, they have never become available in Liberia. Nevertheless, transmission of infections has steadily declined in Liberia similar to other countries[6], likely due to a combination of immunity from infection and vaccination, and circulation of less virulent variants. Despite the present reduced threat from this infection, the initial severe and highly distruptive impact of the SARS-CoV-2 pandemic warns us of the potential spread and consequences of a pathogen against which the global community is unprepared, and the need to assess novel diagnostic tools, particularly for new pathogens, in the population in which they will be used. The scientific and public health community should not assume validated tests will perform similarly in different populations.

## Data Availability

All data produced in the present study are available upon reasonable request to the authors

## Acknowledgements

We thank staff at the Ministry of Health, Liberia and JKF Medical Center for their collaboration in making this study possible. We would also like to acknowledge the contribution of the PCOS study team for implementing the study, and the laboratory staff for conducting the testing. We also gratefully acknowledge all the participants.

## Authors’ Declarations

All named authors have seen and agreed to the submitted version of the paper. The material is original and is unpublished in another journal; it not currently being submitted elsewhere for publication. Preliminary information from the study was presented as an absract at the International Conference of the West African Consortium.

## Declaration of Competing Interest

The authors declare no conflicts of interest.

## Author Contributions

**Moses Badio:** Conceptualization, Data curation, Formal analysis, Investigation, Methodology, Project administration, Supervision, Visualization, Writing – original draft, Writing – review & editing. **Christina Lindan:** Funding acquisition for analysis and writing, Methodology of analysis, Supervision, Visualization, Writing – original draft, Writing – review & editing. **Kumblytee Johnson:** Conceptualization, Data curation, Investigation, Methodology, Project administration, Resources, Supervision, Writing – review & editing. **Cavan Reilly:** Conceptualization, Data curation, Formal analysis, Investigation, Methodology, Visualization, Writing – review & editing. **Julie Blie:** Validation, Data curation, Project administration, Supervision. **Katy Saliba:** Validation, Project administration, Resources, Supervision, Writing – review & editing. **Tamba Fayiah:** Data curation, Writing – review & editing. **John McCullough:** Validation, Data curation, Project administration, Supervision. **James Duworko:** Funding acquisition, Project administration, Resources, Supervision, Writing – review & editing. **Elizabeth Higgs:** Conceptualization, Funding acquisition, Investigation, Methodology, Resources, Supervision, Writing – review & editing. **Jeff Martin:** Funding acquisition, Methodology, Resources, Supervision, Visualization, Writing – original draft, Writing – review & editing. **Lisa Hensley:** Conceptualization, Funding acquisition, Investigation, Methodology, Resources, Supervision, Visualization, Writing – review & editing.

## Notes

**Funding:** This research was supported by the U.S. National Institutes of Health/National Institute of Allergy and Infectious Disease (NIH/NIAID), Partnership for Research on Ebola Virus in Liberia (PREVAIL); the Fogarty International Center/NIAID (U2R TW011281); and the U.S. National Institute of Mental Health/NIH (R25 MH123256).

### Competing Interest Statement

The authors have declared no competing interest.

### Funding Statement

This research was supported by the U.S. National Institutes of Health/National Institute of Allergy and Infectious Disease (NIH/NIAID), Partnership for Research on Ebola Virus in Liberia (PREVAIL); the Fogarty International Center/NIAID (U2R TW011281); and the U.S. National Institute of Mental Health/NIH (R25 MH123256).

### Author Declarations

The National Research Ethics Board of Liberia

